# Global capacity for implementing the Eight Practical Steps to achieve universal access to water, sanitation, and hygiene in healthcare facilities: a qualitative study

**DOI:** 10.1101/2025.09.18.25336017

**Authors:** Darcy M. Anderson, Caralee Sadler, Lucy K. Tantum, Ryan Cronk

## Abstract

Targets to achieve universal access to environmental health services include healthcare facilities. To support improvement efforts, the World Health Organization and UNICEF published the “Eight Practical Steps,” a roadmap for countries to achieve universal access. This document provides clear directions, but many steps are ambitious, and countries’ readiness to complete them is unclear. We conducted a qualitative study to evaluate stakeholder readiness to implement the Eight Practical Steps and documented capacity development needs, using interviews with 45 participants. We applied an implementation science theory—organizational readiness for change—to structure our interviews around factors influencing readiness. Readiness was highest for conducting baseline situation assessments (Step 1), establishing standards (Step 3), improving infrastructure (Step 4), and strengthening the health workforce (Step 6). Readiness was enhanced by stakeholder cooperation, positive attitudes for change, and availability of tools for monitoring, risk assessment, and healthcare worker training. Capacity development was needed to tailor situation assessment tools, training materials, and standards for different contexts. Readiness for change was lower for developing costed roadmaps (Step 2), monitoring progress (Step 5), engaging communities (Step 7), and conducting operational research (Step 8). Stakeholders prioritized earlier steps and had not yet dedicated substantial effort or resources to later ones. Barriers included a lack of tools and guidance. Our results indicate enthusiasm and cooperation to improve environmental health services, but rapid action sometimes came at the expense of strategic planning and learning opportunities. We recommend strengthening funding, personnel, and technical support for planning, monitoring, evaluation, and operational research.

## 1 Introduction

Environmental health services (e.g., water, sanitation, hygiene, waste management, cleaning, and energy) in healthcare facilities are important to protect patients, healthcare workers, and communities (Anderson et al., 2025). They affect individual well-being through increased quality and safety of care and strengthen health systems by improving the usability and functionality of healthcare facilities—including pandemic preparedness and slowed development of antimicrobial resistance (Anderson et al., 2023; Bouzid et al., 2018; Fejfar et al., 2021; Howard et al., 2020; Watson et al., 2019). Despite their importance, access to environmental health services in low- and middle-income countries is poor. In least-developed countries, 32% of healthcare facilities have basic hygiene services, 34% have basic waste management services, 53% have basic water services, and 21% have basic sanitation services (WHO/UNICEF, 2022a).

To encourage progress towards universal access, in 2019, the World Health Organization (WHO) and United Nations Children’s Fund (UNICEF) published *Water, Sanitation and Hygiene in Healthcare Facilities: Practical Steps to Achieve Universal Access to Quality Care* (hereafter the “Eight Practical Steps”) (WHO/UNICEF, 2019). This document outlines a roadmap that national governments and other actors supporting WASH in HCFs can follow to reach universal access targets, focused on high-level national actions that are broken down into eight discrete steps (Table 1).

**Table 1.** Summary of the Eight Practical Steps recommended by the WHO and UNICEF for country action to achieve universal access to environmental health services in healthcare facilities.

| Step | Key actions |
| --- | --- |
| Step 1: Conduct situation analysis and assessment | Assess health and environmental health policies, governance structures, and funding (situation analysis)<br>Determine levels of environmental health services coverage and compliance with relevant policies (assessment) |
| Step 2: Set targets and define roadmap | Define the national approach, intervention areas, and targets<br>Assign roles and responsibilities of stakeholders<br>Develop a budget for environmental health services improvements |
| Step 3: Establish national standards and accountability mechanisms | Define standards that reflect the national context<br>Provide guidance on the design, costing, implementation, and operation of environmental health services<br>Create mechanisms to ensure that facilities meet established standards |
| Step 4: Improve and maintain infrastructure | Construct, rehabilitate, and improve infrastructure to meet standards<br>Develop policies and strategies to operate and maintain infrastructure<br>Ensure that adequate resources are available for operation and maintenance |
| Step 5: Monitor and review data | Establish systems for routine data collection and review<br>Analyze data to measure progress and hold stakeholders accountable |
| Step 6: Develop health workforce | Provide training and mentoring on environmental health services and infection prevention and control practices |
| Step 7: Engage communities | Include communities in developing policies and ongoing review of data on environmental health services coverage and implementation<br>Coordinate with community environmental health service providers where appropriate |
| Step 8: Conduct operational research and share learning | Document approaches, challenges, and solutions<br>Reflect on and revise programmatic strategies based on external review and research<br>Assess the potential for scale-up of innovative approaches |

Countries can voluntarily report progress on seven of the eight steps to a database maintained by the WHO and UNICEF using a stoplight system ranging from “no progress” to “completed/achieved” (WHO/UNICEF, n.d.). As of the end of 2024, most countries reported having started each step in some capacity—or having plans to start—but none had completed all steps. The steps with the most progress were Step 1 for conducting baseline assessments and Step 3 for establishing national standards, with the greatest progress specifically on waste management standards.

Key barriers to progress are likely the complexity of each step, lack of specific guidance on how to complete it, and a need for capacity development. The Eight Practical Steps require coordination and action by many actors—often across multiple sectors at the national and sub-national levels. The Eight Practical Steps framework provides a broad roadmap but little specific guidance. Other companion documents released by the WHO and UNICEF (e.g., the 2024 *Global Framework for Action* (WHO/UNICEF, 2024a)) reaffirm the importance of the Eight Practical Steps and provide additional recommendations. However, targets are ambitious (e.g., 100% of least developed countries achieving basic environmental health services in all healthcare facilities by 2030), and the recommendations to achieve them are similarly broad. For example, the WHO and UNICEF recommend that countries “integrate [environmental health services] into health system planning, programming, financing, implementation and monitoring,” but how countries can accomplish this remains unclear.

Understanding stakeholders’ capacity to implement the Eight Practical Steps can help inform more specific and actionable recommendations and guide the development of tools, training, and other resources to support implementation. There are currently no assessments to evaluate readiness to implement the Eight Practical Steps, despite its importance in policy frameworks promoted by the WHO and UNICEF (WHO/UNICEF, 2024a).

We conducted an in-depth qualitative study of stakeholder capacity and readiness to implement the Eight Practical Steps, using a theory from implementation science called the “organizational readiness for change,” which identifies specific drivers of readiness to implement complex interventions and policies (Weiner, 2009). Our objectives were to (1) evaluate stakeholders’ readiness for the Eight Practical Steps and (2) document perceived capacity development needs to increase readiness.

## 2 Methods

### 2.1 Conceptual framework

We applied a theoretical framework from implementation science to assess readiness for implementing the Eight Practical Steps: Weiner’s theory of organizational readiness for change (Weiner, 2009). This theory was developed to assess readiness to implement and sustain organizational changes and to prepare for implementing evidence-based health projects and programs. The theory proposes that readiness for change is driven by willingness (e.g., whether stakeholders believe the change is important, necessary, or beneficial) and ability (e.g., actual and perceived resource availability, time and effort to implement the change, favorability of the political climate). Supplemental File 1 provides a more extensive breakdown of specific drivers related to willingness and ability.

To evaluate stakeholder readiness to implement the Eight Practical Steps, we examined the extent to which these drivers related to willingness and ability to implement change were present versus absent. To evaluate capacity development needs, we assessed gaps in readiness drivers and documented specific actions that stakeholders perceived would increase their ability or willingness to implement the Eight Practical Steps.

### 2.2 Participant recruitment and sampling

To develop our initial sampling frame, we reviewed resources from 2022 and 2023 on washinhcf.org (the most extensive online repository of resources on environmental health services in healthcare facilities) to identify individuals and organizations working on environmental health services in low- and middle-income countries. We prioritized individuals in senior positions, such as project leads or training specialists. We also contacted the WHO and UNICEF to identify additional individuals who were highly influential or knowledgeable about implementation and capacity building. We invited all individuals in this initial sampling frame to participate in interviews.

We then employed a snowball sampling approach and asked those participants to nominate a colleague (ideally from a different organization or with a different role) for a second round of recruitment. After this second round of interviewing, we reached saturation and ended recruitment.

### 2.3 Data collection

We collected data using semi-structured qualitative interviews. We developed an interview guide that was divided into sections to evaluate each of the Eight Practical Steps. For each step, we asked about current implementation activities, roles and responsibilities, available tools, resources, and technical support, and perceived capacity development needs.

A team comprising one lead postdoctoral researcher and five Masters of Public Health students conducted interviews from June through August 2023. One person conducted each interview in English or French using Zoom video conferencing. Interviews lasted approximately 40 minutes to one hour. We used Zoom’s built-in audio recording and transcription feature to capture transcripts. We spot-checked ten transcripts (27%) for accuracy and identified only a small number of errors. In all cases, errors were minor misspellings or word replacements that did not affect the interpretation of the transcripts. We, therefore, proceeded directly to use the Zoom transcripts for analysis, addressing misspellings and other minor errors as necessary during coding.

For participants who wanted to participate in the study but could not join a real-time Zoom interview, we supplied an abbreviated questionnaire for written responses. The most common reasons for choosing the written alternative were language barriers, low internet bandwidth, or scheduling challenges across time zones.

### 2.4 Analysis

We created a codebook containing constructs for readiness drivers based on the theory of organizational readiness for change (Weiner, 2009)One author (CS) applied the codebook to ten transcripts. Then, two authors (DA and CS) convened to review and revise the codebook to ensure that definitions captured relevant findings and that codes were applied consistently. One author (CS) then coded the entire dataset using NVivo qualitative analysis software.

We co-coded each relevant section of text with a code for readiness determinants and a code for one or more of the Eight Practical Steps. We used the coding matrix query function in NVivo to stratify the readiness determinants under each Eight Practical Steps. Two authors analyzed the resulting data to assess trends in the readiness codes that appeared under each of the Eight Practical Steps.

### 2.5 Ethics

This study was designated as not human subjects research by the Institutional Review Board of the University of North Carolina at Chapel Hill (IRB #22-3235). We obtained verbal informed consent from all interview participants.

## 3 Results

### 3.1 Participant demographics

We interviewed 37 participants and received six written responses. Most participants (67%) were from the WHO African region; 37% worked for non-governmental organizations, 23% for multilateral organizations, and 21% for government agencies. Most (77%) had worked for their current employer for at least five years (Table 2).

**Table 2.** Demographics of interview participants (n=43).

| Participant characteristic | N (%) |
| --- | --- |
| *WHO region* |  |
| African Region | 29 (67%) |
| South-East Asian Region | 6 (14%) |
| Region of the Americas | 3 (7%) |
| European Region | 2 (5%) |
| Western Pacific Region | 2 (5%) |
| Eastern Mediterranean Region | 1 (2%) |
| *Organizational affiliation* |  |
| Non-governmental organization | 15 (35%) |
| Government | 10 (23%) |
| Multilateral organization | 9 (21%) |
| Academic institution | 5 (12%) |
| Private sector | 2 (5%) |
| Donor agency | 2 (5%) |
| *Years of experience in current position* |  |
| 0-2 | 14 (33%) |
| 3-5 | 14 (33%) |
| 6-10 | 11 (26%) |
| More than 10 | 4 (9%) |
| *Years of experience in the current organization* |  |
| 0-2 | 6 (14%) |
| 3-5 | 4 (9%) |
| 6-10 | 15 (35%) |
| More than 10 | 18 (42%) |

### 3.2

### 3.3 Organizational readiness and capacity development needs

We found that readiness was higher for steps to perform baseline situation assessments, establish national standards, improve infrastructure, and develop the health workforce (Steps 1, 3, 4, and 6). Driving readiness for these steps was higher willingness and perceived ability— partly due to higher importance and urgency placed on these steps, lower perceived complexity, and wider availability of tools. Readiness was lower for steps to set targets and create costed roadmaps, monitor data, engage communities, and conduct operational research (Steps 2, 5, 7, and 8). Readiness for these steps was hindered primarily by lower perceived ability because stakeholders described a lack of resources and tools to implement these steps. Stakeholders believed these steps were important, but willingness to complete them was lower because they were perceived to compete for time and resources. Steps 1, 3, 4, and 6 were a higher priority.

Table 3 summarizes key readiness drivers and capacity development needs for each step. However, readiness was not uniform across steps. For example, readiness for Step 2 was high for setting targets but low for developing costed roadmaps. We disaggregate findings across specific components of each step in our narrative summary where relevant.

**Table 3.** Key readiness factors and capacity development needs for the Eight Practical Steps among environmental health services stakeholders.

| Step | Key readiness factors | Capacity development needs |
| --- | --- | --- |
| Step 1: Conduct situation analysis and assessment | **Willingness.** Participants recognized the importance of assessment, but facility-level staff were sometimes skeptical. Many organizations had already begun baseline assessments, with collaboration between partners strengthening readiness; few had performed situation analysis.<br>**Perceived ability:** Assessment tools are available, but some participants perceived existing tools as insufficiently detailed or unsuited for their context. Assessments were resource-intensive, and limited staff, time, and funding availability hindered readiness. | Contextualized, digitized assessment tools<br>Guidance and more detailed situation analysis tools |
| Step 2: Set targets and define roadmap | **Willingness:** Target-setting was perceived as important for change. Stakeholders believed that other stakeholders support change, creating a collaborative, supportive context.<br>**Perceived ability:** Limited experience with costing, lack of cost data, and uncertainty about future needs posed constraints. Stakeholders lacked clear examples and templates. | User-friendly costing tools<br>Training for the budgeting and costing process<br>Templates and examples for developing costed roadmaps |
| Step 3: Establish national standards and accountability mechanisms | **Willingness:** High levels of enthusiasm for action across government and non-government actors increased readiness. Participants identified and supported government “champions” to lead change.<br>**Perceived ability:** Governments increasingly had staff positions to create and enforce standards. Developing continuity of partnerships and champions was key for readiness. | Contextualized policies, regulations, and standards at the national level<br>Government partnerships and staffing |
| Step 4: Improve and maintain infrastructure | **Willingness:** Participants expressed enthusiasm for implementing infrastructure improvements and placed high value on this step compared to others. | Financial resources for constructing and maintaining infrastructure (e.g., cleaning, monitoring, repairing) |
|  | **Perceived ability:** Strong planning skills to prioritize and implement improvements increased readiness. The primary barrier to implementation was lack of funds. Readiness to operate and maintain infrastructure after construction was less, in part due to lack of trained staff. | Recruiting and training staff on operations and maintenance |
| Step 5: Monitor and review data | **Willingness:** Participants expressed concerns about sharing data but were more willing to collaborate if data would be used to improve services.<br>**Perceived ability:** Data dashboards informed planning and motivated change. | Tools to collect and share data<br>Guidance to adapt baseline assessment tools for routine monitoring<br>Staff and financial resources to conduct monitoring |
| Step 6: Develop health workforce | **Willingness:** There was both high demand for trainings and collaboration between organizations to establish master trainer networks to meet this demand.<br>**Perceived ability.** Organizations were confident in their ability to access trainers, identify tools, and set up training systems. However, readiness to meet the full demand was low, as training was time-intensive, and the need for refresher training posed a burden for organizations. Digital tools alleviated some logistical difficulties.<br>Training tools for clinical staff were widely available, which increased readiness. | Staffing and resources for delivering regular refresher training<br>Training tools tailored to country-specific contexts and additional languages<br>Training tools for non-clinical staff (e.g., maintenance technicians) |
| Step 7: Engage communities | **Willingness:** Participants were interested in developing strategies to engage community members more effectively.<br>**Perceived ability:** Organizations lacked dedicated funds, plans, and guidance for community engagement. While some performed | Tools and guidelines for planning and conducting outreach<br>Communication materials |
|  | outreach, efforts were not consistently monitored. |  |
| Step 8: Conduct operational research and share learning | **Willingness:** Operational research was perceived as a lower priority compared to other steps.<br>**Perceived ability:** Participants had lower confidence in their ability to conduct operational research and reported a lack of financial resources and research training. | Financial resources<br>Academic partnerships and/or staff training to conduct research |

#### 3.3.1 Step 1: Conduct situation analysis and assessment

Both willingness and perceived ability were high for Step 1. Participants primarily described readiness for baseline assessments (i.e., one-half of Step 1 for assessing environmental infrastructure access and related behaviors). Most participants reported that baseline assessments were completed or in progress. Participants rarely described assessing policies, governance structures, or other factors that would be included in situation analysis (i.e., the other half of Step 1).

High perceived ability was strongly influenced by the widespread availability of tools to assess the baseline status of environmental health services. Most participants used the Water and Sanitation for Health Facility Improvement Tool (WASH FIT), though some described others. WASH FIT was preferred because it was perceived as easy to use and packaged with digitized indicator banks and training. However, some participants noted that WASH FIT did not necessarily yield enough depth of information:

> *[WASH FIT] is a tool that I think is meant to be used by anybody, which I think is one of the strengths of the tool. It doesn’t take necessarily a WASH expert to fill it in. It doesn’t take an engineering expert to fill it in. So while it’s great at giving that broad understanding, I think once you get past that, you kind of want a little bit more in depth. Like, okay, so what are the issues with the water? And I always kind of found that that was something I had to chase around a little bit and get a little bit more in depth knowledge. -Technical officer, multilateral organization*

Staff, time, and funding for assessments were limited and hindered readiness. Participants noted that assessments required substantial resources to complete. Isolated and rural healthcare facilities required comparatively more resources for assessments, and resource limitations influenced where assessments could be completed. Collaboration between stakeholders helped mitigate resource barriers and enhanced readiness (e.g., sharing assessment data and delegating assessment areas to different partners). There was also strong support from multilateral and bilateral agencies and international non-governmental organizations (NGOs) for training national and subnational teams on assessment methods.

Participants noted a need to adopt digitized tools and adapt them to the local context. Manual data entry was time intensive, and participants suggested using electronic versions to help with ongoing monitoring and data sharing. Participants also suggested developing tools adapted to specific contexts (e.g., different languages, sizes and types of healthcare facilities, local capacities).

Willingness to complete Step 1 was generally but not universally high. Many participants described situation assessments favorably and perceived them as important to prepare for other steps. Some participants described how data was leveraged for advocacy purposes, particularly communicating with health sector stakeholders who were less familiar with environmental issues. However, participants also reported that on-the-ground implementers and healthcare facility staff were sometimes skeptical of assessments, seeing them as a waste of resources that could be better directed toward building infrastructure.

#### 3.3.2 Step 2: Set targets and define roadmap

Participants described high willingness and perceived ability to set targets for improving environmental health services. Attitudes towards change were favorable, with participants valuing environmental health services and supporting ambitious targets to improve them. Most participants perceived that other stakeholders valued change, including growing awareness and support among non-environmental health sector stakeholders and policymakers.

Willingness to develop costed roadmaps was high, but perceived ability was low. Participants described how their countries wanted to develop costed roadmaps but lacked relevant experience. One participant from a multilateral agency described a lack of good examples as a barrier to readiness:

> *We’ve had a few countries ask us for a template to work on [roadmaps or action plans]. We don’t really have one. There have not been that many good examples of country roadmaps that we can use as a template. -Technical adviser, multilateral organization*

A commonly described barrier to readiness was a lack of robust cost data. Some participants used estimates from cost data from prior projects and programs to estimate costs. However, these were from a small geographic area and homogenous set of healthcare facilities, raising questions about the accuracy of extrapolating these data to the national level. Participants recommended capacity development for more user-friendly tools for creating budget plans and greater availability of cost data.

Even when costs were known, uncertainty about resource availability was also a barrier to planning. Targets were often ambitious beyond currently available resources, but there was hope of future funding increases. Several participants identified a capacity development need to help stakeholders identify funding sources and advocacy for funding alongside costed roadmaps, including one participant who stated:

> *[I would like] to have a tool that can help them [NGO country offices] to practically identify resources. This could be finances; this could be to do that kind of assessment that within their own, environment setup they can identify. Resources with which they can plan to move forward. But at the same time also, tools. Which for every domain, for example, we need to come up with the tools that [are] more practical. -Regional WASH advisor, multilateral organization*

#### 3.3.3 Step 3: Establish national standards and accountability mechanisms

Readiness for Step 3 was like that of Step 2. Participants described enthusiasm for action and strong collaboration between country governments and development partners (i.e., high willingness) but sometimes a lack of data and detailed technical guidance (lower perceived ability). The availability of guidelines from the WHO and country governments enhanced readiness, as these could be used as templates and adapted. Multilateral agencies—notably WHO and UNICEF—worked closely with government agencies to develop national standards. However, participants noted that standards did not always account for the nuanced needs of different subnational regions and facility types, and they described this as an area for capacity development.

Readiness was enhanced by development partner collaboration that supported government-led action. Participants described increased staff positions within national and local governments to promote and enforce standards. NGOs strengthened readiness by consolidating their efforts around government leadership:

> *Whatever we do in [our country] is fully aligned with the national priorities. And when we say national priorities, we actually work closely with the government… We actually go to visit them. We, we sit with them… We actually ask them, ‘Where [do you] you want us to work?’ -Regional WASH advisor, non-governmental organization*

Other NGO participants noted that they had played an active role in identifying and supporting champions within Ministries of Health and local governments. One NGO participant described multi-year efforts to develop partnerships and support champions within government to promote the uptake of policies and standards:

> *We noticed that we needed policy approval, so we started developing key champions and over the course of the last 4 to 5 years [we] developed strong partnerships and champions within the Ministries of Health. And so, they are the ones that are now leading*… *-Program officer, non-governmental organization*

#### 3.3.4 Step 4: Improve and maintain infrastructure

Participants described a strong willingness and ability to implement infrastructure in most contexts. Stakeholders described a strong ability to plan and prioritize infrastructure improvements, aided in some cases by structured decision-making tools, specifically WASH FIT. Generally, the choice of solutions and how to implement them was seen as straightforward and “obvious,” with the primary implementation barrier being lack of funding. One participant from a donor foundation described how their grantees would prioritize actions and create contingency plans based on resourcing:

> *The challenge there is always now try to mobilize resources… I almost feel like there’s one [plan] that they can’t afford and one they can really do… Or maybe an attainable wish list and then the third tier one is, like, ‘If we ever get a money, this is what we want to do*.*’ I feel like that’s the way that a lot of our grantee partners end up once they have the action plan, because they can’t probably do all of it. -Program officer, donor organization*

The perceived ability to implement improvements was notably lower in contexts where healthcare facilities lacked access to basic water services. This negatively influenced willingness to attempt infrastructure improvement for environmental health services besides water. One participant described how they were demotivated to act because they believed that attempts to improve cleaning, hygiene, or other environmental services would fail without water:

> *A lot of times the complaint is like, ‘Well, we can’t do anything. We don’t have water,’ and it’s completely unmotivating, and so a big piece of the work was trying to find ways to motivate. -Technical officer, multilateral organization*

Across all contexts, readiness to maintain improvements was lower than readiness for initial implementation. Some participants described how resources were focused on constructing new infrastructure, and many participants reported a capacity development need to dedicate resources and hire and train more staff to clean, repair, and maintain infrastructure. Multiple participants specifically spoke about training for cleaners and believed that environmental tasks should not be placed on healthcare providers.

### 3.3.5 Step 5: Monitor and review data

Readiness for Step 5 was lower than that of Step 1, even though both require collecting data on environmental health services coverage. The perceived ability was similar—largely because the same tools for baseline assessments in Step 1 could be used for Step 5. However, participants described using WASH FIT and other assessment tools to conduct baseline assessments but reported that the same tools were not necessarily systematized into sustained monitoring.

This was partly due to a lower willingness to establish monitoring systems due to concern over how they may be used. On one hand, participants recognized that monitoring data were important for tracking progress, holding stakeholders accountable, and recognizing and rewarding progress. However, they feared monitoring data could be used punitively when it showed poor conditions.

> *One of the things that I think has been discouraging people is that monitoring has been seen coming from someone where some somebody comes to monitor—not for improvement, but for checking on what is wrong, what’s not being done. And in any way that is punitive. Sometimes it goes back as punishments to the implementers. But in this case, for us, we say… [that] if monitoring can be used as something that is cherished by the very owners or the people were implementing, that it motivates them to see their level of improvement. -Regional WASH adviser, non-governmental organization*

Participants described ways to increase willingness to engage in monitoring. For example, when monitoring was linked to mechanisms to provide support in response to lapses in service quality, there was also greater trust and willingness to participate.

Developing dashboards to display data for decision-making purposes was a commonly described capacity development need, though there were concerns about how that data would be viewed and used. Some participants also perceived concerns among government regulators at the national and subnational levels, where there was a desire to have access to monitoring data but reluctance to share that data with others. One proposed solution to this was to develop data dashboards exclusively for internal use:

> *Where we saw the sensitivity of sharing that data, the latest thing that we’re looking at is: Can we have just country sites where ministries of health can log on and see their own data that not necessarily everyone else sees until they’re comfortable with sharing it, but at least to start getting some kind of an aggregation of data and some kind of an overview—whether it’s a district level, province, level or national level, depending on the roll out of WASH FIT. Just to be ready with those data collection tools and dashboards, and to facilitate that decision-making. -WASH advisor, non-governmental organization*

#### 3.3.6 Step 6: Develop health workforce

Readiness to conduct Step 6 was high, both in terms of willingness and perceived ability. Many participants described a high demand for training, as well as a corresponding supply of master trainers, mentors, role models, and established systems to deploy these individuals to deliver training. Participants described having content available and ready to deliver trainings to cleaners and healthcare workers, noting that the COVID-19 pandemic had increased the availability of training materials, particularly related to hygiene and infection prevention and control.

The primary barrier to readiness was the perceived ability to manage the logistics of delivering established training materials. Participants described that training was time intensive, and it was difficult to host enough training to cover all staff. The need for recurring and refresher training placed additional demand on training systems that were perceived as already overburdened. An additional challenge was that healthcare workers had to take time off from clinical duties to attend training, which contributed to staffing challenges:

> *The healthcare providers themselves may not attend their training a hundred percent. You cannot keep, for example, work at the hospital stalling, yet there are people waiting in their queues. Some will attend the training, while others attending to their clients. -Project officer, non-governmental organization*

The availability of digital tools mitigated this barrier and enhanced readiness. Digital training materials could be disseminated more easily, and content could be consumed asynchronously. One participant described establishing a network of champions to disseminate digital materials:

> *After [champions] were selected, the first thing was they were all added to a WhatsApp group…. So the concept was we would send them those materials and then they were supposed to forward them to all their clinical groups, WhatsApp groups. So we have nurses and doctors and admins and they were supposed to then forward them to their department WhatsApp groups…. So, not a perfect distribution chain, but—just given their hospitals run 24-hours a day—we couldn’t possibly train everybody in person. So it seemed like the best distribution approach. -Epidemiologist, academic institution*

Multiple participants recommended increasing the availability and type of content covered by online, asynchronous training materials as a capacity development need. Participants also described a need to expand the language selection of available training materials and tailor them to countries’ specific national guidelines and facilities’ standard operating procedures.

Few participants explicitly described willingness or ability for workforce development beyond training healthcare facility staff beyond healthcare workers and cleaners. One participant remarked on this gap and specifically called for a need to hire and train administrative staff and technicians that supported environmental health services outside patient care settings:

> *There’s no capacity building for logisticians or for engineers, and people don’t know how to repair things. What’s missing is sometimes is engineers, but more frequently… qualified technicians, rather than engineers. People that will be hands on, that are able to maintain a system. Otherwise you’re asking healthcare workers and providers to just do too much, and things end up like getting missed. It’s not fair to ask them to do everything. -WASH specialist, non-governmental organization*

#### 3.3.7 Step 7: Engage communities

Steps 7 and 8 displayed the lowest readiness of all steps. Many participants reported no current or prior efforts for community engagement. However, one regional WASH officer for a multilateral organization reported that community engagement was difficult to track because it tended to be grassroots and not reported through national monitoring systems.

While uncommon, most participants reported a high value in engaging community members. There was a shared desire among participants to increase engagement efforts. As one participant described:

> *You cannot to do WASH in a healthcare facility without the community. You won’t go far. -WASH program officer, non-governmental organization*

A barrier to readiness was that organizations lacked no dedicated funds or specific planned activities. Readiness was also impeded by a lack of specific guidance on how to engage communities or what exactly comprised community engagement. For participants who reported some form of engagement, there was a wide range of activities. Some reported that community members contributing funds or labor to constructing infrastructure was engagement. Others described community engagement as visits by health promotion officers or community health workers to do education. We found some reports of including community members in healthcare facility WASH or infection prevention committees, but participants reported that these community representatives did not necessarily receive training or capacity building to help them meaningfully contribute.

Participants suggested capacity development needs for outreach tools, guidelines on how to involve communities, and tools and training around context-appropriate communication with lay community members.

#### 3.3.8 Step 8: Conduct operational research and share learning

Along with Step 7, Step 8 overall displayed the least readiness of all the Eight Practical Steps. Most participants stated that they were doing little to no operational research. Some participants indicated a desire to do operational research but believed that their organizations lacked the appropriate training or expertise. These individuals identified a capacity development need to form partnerships with academic institutions to conduct operational research and/or train local staff. Other participants reported that operational research was not a current priority. In these cases, participants did not perceive an urgent capacity development need, preferring to focus efforts on earlier steps first and to delay operational research until prior steps were completed or at least well underway.

## 4 Discussion

We assessed stakeholders’ readiness and documented perceived capacity development needs to increase readiness to implement the Eight Practical Steps framework (WHO/UNICEF, 2019). The Eight Practical Steps are an important policy framework developed and promoted by the WHO and UNICEF to achieve universal access to environmental health services in healthcare facilities by 2030. Understanding stakeholders’ readiness is critical to strengthening and supporting implementation efforts.

We found higher readiness for Steps 1, 3, 4, and 6. The driving factors for these steps were both higher willingness and perceived ability, in part due to the higher importance and urgency placed on them, lower perceived complexity, and wider availability of tools. Readiness was lower for Steps 2, 5, 7, and 8; stakeholders were willing to complete these steps but perceived lower urgency and fewer tools and resources to support implementation.

Our findings broadly align with data that countries self-report into a progress tracker maintained by the WHO and UNICEF (WHO/UNICEF, n.d.). The proportion of countries that have completed a given step is highest for Step 1 (52% for baseline assessments, 40% for situation analyses) and Step 3 (59% for waste management standards, 45% for water, sanitation, and hygiene standards). No more than 20% of countries report completing any other step. Steps 2, 4, 5, 6, and 7 have many countries that have initiated but not completed the step. Step 8 is not monitored. Progress data tracked by WHO and UNICEF is consistent with our findings that readiness varies across specific actions under each step (e.g., baseline assessments versus situation analysis under Step 1). Steps with a high proportion of initiation but a low proportion of completion also align with our findings that stakeholders had a high willingness to complete all of the steps but perceived a lack of ability, particularly for Steps 2, 5, 7, and 8. Taking action to initiate change is a key readiness signal but is not sufficient to fully implement a change without additional factors that support stakeholders’ ability to implement (Castañeda et al., 2012; Weiner, 2009; Weiner et al., 2008).

The most cited readiness driver related to the perceived ability to implement the Eight Practical Steps was the availability of tools and resources to guide and support specific actions under each step. Correspondingly, steps with lower readiness frequently cited developing these resources as a capacity development need. Stakeholders’ responses are consistent with reviews of available literature on available tools and resources.

Steps 1 and 5 have well-established mechanisms for monitoring and assessment, using indicators for tracking the Sustainable Development Goals (WHO/UNICEF, 2018) and assessment tools contained in WASH FIT (WHO/UNICEF, 2022b). For Step 2, some resources exist for costing and budgeting (Anderson, Cronk, et al., 2021; Anderson et al., 2020; Anderson, Tracy, et al., 2021). Some efforts have been made to leverage these tools for policy change at the local level (Chettry et al., 2024), but how cost data will translate to nationally costed roadmaps remains unclear. For Step 3, some technical guidelines exist to support countries in setting national standards, but the most comprehensive (Adams et al., 2008) has not been updated in over a decade. More recent individual guidelines exist for waste management (Chartier, 2014); water, sanitation, and hygiene do not have dedicated WHO guidelines but are addressed as components of other guidelines (e.g., infection prevention and control (WHO, 2016). For Step 4, systematic reviews have examined the effects of environmental health services on selected outcomes (e.g., healthcare-acquired infections (Watson et al., 2019) and care seeking (Bouzid et al., 2018). Still, no comprehensive efforts have been conducted to compile interventions and guide selection. For Step 6, systems for training medical professionals have incorporated aspects of environmental health services—particularly hygiene—into infection prevention and control curricula for decades. Cleaners and waste handlers have historically been neglected, though resources have become available more recently (The Soapbox Collaborative, 2018; WHO, 2024). Workforce development is notably neglected as it relates to maintenance, supply chain management and logistics, and other non-clinical roles. It also likely contributes to low readiness observed in other steps, such as Step 2 for costed roadmaps.

To our knowledge, there are no tailored guidelines on how to conduct community engagement for Step 7 or operational research for Step 8 specifically related to environmental health in healthcare facilities. General toolkits exist (see, e.g., (WHO, 2020)) but were not well known to participants in our sample—nor do we suspect they are broadly familiar to other stakeholders beyond our sample.

While willingness to change and availability of tools were key readiness drivers, they were insufficient. While stakeholders believe that baseline assessments for Step 1 and monitoring for Step 5 were both important—and ostensibly, the same tools could be used for both—there was a notable gap in readiness. We suspect several factors contribute to this gap. First, is the availability of financial and human resources. One-off assessments are logistically simpler to execute and do not require intensive investment to develop and sustain nation-wide reporting systems. Second, we hypothesize that stakeholders hold an implicit preference for steps that yield or strongly support rapid, observable increases in infrastructure coverage. Steps related to monitoring, evaluation, and learning are perceived to be in competition for the resources needed for infrastructure construction or other more direct intervention implementation efforts. Multiple stakeholders described how Step 5 (monitoring) and Step 8 (operational research) were important but could wait until goals for increasing access were achieved. This suggests that stakeholders view these steps to be in competition, rather than working in alignment towards the same goal.

### 4.1 Implications for policy and practice

This higher readiness for earlier steps likely signals future sustainability challenges and missed opportunities for learning and improvement. Evidence suggests that environmental health services interventions struggle with underperformance and sustainability when there is a strong emphasis is placed on rapid implementation without corresponding efforts to understand and integrate into local systems and context and to monitor, evaluate, and incentivize their continued performance (Abu et al., 2021; Isunju et al., 2022; Tantum et al., 2024).

Performance and sustainability challenges are by no means unique to environmental health services in healthcare facilities and have been well-documented in community-based water, sanitation, and hygiene programming for decades (Martin et al., 2018). More recently, healthcare facilities have only explicitly been recognized as a priority setting—being included in international monitoring efforts for the first time under the Sustainable Development Goals in 2015. Nevertheless, early evidence suggests similar challenges. For example, WASH FIT is one of the most popular programmatic approaches for environmental health services in healthcare facilities, and systematic reviews and narrative reports find challenges with sustainability despite its emphasis on continuous quality improvement techniques (Kpodzro et al., 2024; Lineberger et al., 2024; WHO/UNICEF, 2024b).

These early signs offer an opportunity to correct the course. Within the Eight Practical Steps framework, steps to conduct operational research, engage communities, establish monitoring systems, and train the workforce can all reasonably be expected to support sustainable, systematic change. Furthermore, we found that stakeholders generally report a willingness to engage in these steps but perceive a lack of ability to implement them.

One major action that could be taken is building capacity for Step 5 around monitoring—both high-level national monitoring of progress towards universal access targets and creating systems that can track performance and sustainability at a more granular program level. For example, stakeholders noted a capacity development need to develop tailored, digitized systems monitoring. This will likely require support and investment from donors and research partners to develop and political will from policymakers to implement. Well-developed indicators already existing under tools such as WASH FIT improve readiness, though additional resources will be needed. WASH FIT and other indicators focus on outputs such as infrastructure access; there are fewer tools for measuring inputs or longer-term outcomes and impacts (Lineberger et al., 2024)Efforts to develop more comprehensive monitoring tools for programmatic use and identify a streamlined set of indicators that can be feasibly integrated into high-level national monitoring would support efforts to implement the Eight Practical Steps.

Another critical opportunity to strengthen capacity is integrating Step 8 for operational research into other steps as a concurrent rather than separate activity. Stakeholders in our sample perceived research as a complex and challenging process that required academic partnerships, and they often opined that research should be conducted as a standalone evaluation activity after intervention delivery.

While common, we argue that this perception creates several unnecessary challenges and that correcting it could enhance stakeholders’ readiness to implement the Eight Practical Steps, not just Step 8, for operational research. At its core, operational research is intended to integrate into programs and create actionable findings that improve implementation and address real-world problems for practitioners (Bauer & Kirchner, 2020; Eccles & Mittman, 2006; Theobald et al., 2018). As a subset of implementation science, operational research is grounded in the principles that research should be driven by the needs and priorities of practitioners and be adapted to fit within programmatic activities (Monks, 2016). Partnerships with academic institutions may strengthen operational research but are not strictly necessary.

Another key feature of operational research is its focus on understanding implementation processes and identifying opportunities to improve them (Bauer & Kirchner, 2020; Eccles & Mittman, 2006; Monks, 2016; Theobald et al., 2018). This is directly relevant to strengthening the capacity to implement the Eight Practical Steps. Stakeholders in our sample identified a capacity development need for templates, examples, and guidelines on developing costed roadmaps (Step 2). Operational research can develop these resources. For example, a 2024 operational research study in Nepal documented the process of budgeting and advocacy project (Chettry et al., 2024). The project led to policy change at the municipal level to establish a fund for the operation and maintenance of environmental health services. The focus of the research was documenting the activities of the project and its successes and challenges such that it could be replicated elsewhere. Notably, the methods used were predominantly qualitative and narrative documenting project activities, which are techniques that should be familiar and achievable for most practitioners.

Operational research efforts like this can and should be used to support implementing the Eight Practical Steps framework. Timeliness of research findings is important for their useful translation into practice (Cash et al., 2003). Delaying operational research until after other steps are complete dilutes its usefulness and misses opportunities to learn and improve in real-time. Efforts to train practitioners to integrate monitoring and learning as a form of operational research into their day-to-day activities can support implementation and sustainability of efforts to reach universal access. Disseminating findings from this operational research so that they are readily accessible to stakeholders beyond the immediate project team will also be critical to building global capacity.

### 4.2 Limitations

Our sample comprised individuals working predominantly at the national or international level in leadership and management roles, who were accessible for interventions by Zoom in English and French. Our sample did not include individuals working at the local levels, who we expect may have different perspectives on readiness and capacity development needs to implement the Eight Practical Steps. Future research that samples multiple levels of stakeholders from the national to the local level within specific country case studies would be valuable.

## 5 Conclusions

We evaluated readiness to implement the Eight Practical Steps and found that readiness was higher for earlier steps related to conducting baseline assessments, setting targets, and implementing interventions to improve infrastructure and train the health workforce. While stakeholders perceived all steps to be important, they were deprioritized partly because they were seen as less urgent and because they perceived fewer resources and tools to implement them. Existing tools and resources centered around assessing baseline conditions to implement interventions, with stakeholders identifying capacity development needs to establish systems for monitoring, evaluation, and learning, and other forms of operational research that may increase performance and sustainability.

Our results indicate a promising enthusiasm and cooperative spirit to improve environmental health services in healthcare facilities, but an emphasis on rapid action sometimes at the expense of strategic planning and missed opportunities for learning and improvement. To ensure that enthusiasm and resources are effectively directed, we recommend that international efforts and country-level programs strengthen funding, personnel, and technical support for planning, monitoring and evaluation, and operational research and learning, which is conducted concurrently with efforts to implement interventions and train the health workforce.

## Data Availability

All data produced in the present study are available upon reasonable request to the authors

## 6 Acknowledgements

We thank the following people who helped conduct interviews for this study: Anushka Banerjee, Doreen Kaphalanya, Amelia Martin, Montana Scott, and Brooklyn Story. We also thank the interview participants who generously donated their time and expertise to this study. This work was funded in part by the Wallace Genetic Foundation. LKT was supported by the National Science Foundation Graduate Research Fellowship under Grant No. DGE-2040435. DMA was supported by a grant from the Unite States National Institutes of Environmental Health (T32ES007018).

